# COVID-19 Trend and Forecast in India: A Joinpoint Regression Analysis

**DOI:** 10.1101/2020.05.26.20113399

**Authors:** Aalok Ranjan Chaurasia

**Affiliations:** MLC Foundation

**Keywords:** COVID-19, Pandemic, India, Trend, Forecast, Joinpoint Regression

## Abstract

This paper analyses the trend in daily reported confirmed cases of COVID-19 in India using joinpoint regression analysis. The analysis reveals that there has been little impact of the nation-wide lockdown and subsequent extension on the progress of the COVID-19 pandemic in the country and there is no empirical evidence to suggest that relaxations under the third and the fourth phase of the lockdown have resulted in a spike in the reported confirmed cases. The analysis also suggests that if the current trend continues, in the immediate future, then the daily reported confirmed cases of COVID-19 in the country is likely to increase to 21 thousand by 15 June 2020 whereas the total number of confirmed cases of COVID-19 will increase to around 422 thousand. The analysis calls for a population-wide testing approach to check the increase in the reported confirmed cases of COVID-19.

## Background

Total number of reported confirmed cases of COVID-19 in India crossed 100 thousand mark on 19 May 2010 according to the database maintained by the World Health Organization. The first confirmed COVID-19 case in India was reported on 30 January 2020 but no confirmed COVID-19 case was reported during 4 February 2020 through 1 March 2020. By 15 March 2020, more than 100 confirmed cases of COVID-19 were reported which increased to 500 by 24 March 2020 when the nation-wide lockdown was announced in the country. Since then, the number of daily reported confirmed cases crossed the 10000 mark by 14 April 2020 and the 50000 mark by 7 May 2020. The progress of the disease presents a challenge for data scientists to model it as the epidemiological characteristics of the disease are yet to be fully explained. There is currently no treatment for COVID-19 and no vaccine is available to protect from novel coronavirus. The uncertainty about the progress of COVID-19 pandemic, therefore creates additional pressure on the epidemiologists and public health experts on how to control it. In such a situation, forecasting COVID-19 cases is very important for planning and implementing infection containment and pandemic control measures. In this context, an analysis of the trend in the daily reported confirmed cases of COVID-19 may provide useful information about how the COVID-19 pandemic has progressed in India. The trend analysis also permits forecasting the likely trend in the reported confirmed cases of COVID-19 in the immediate future.

A trend analysis of daily reported confirmed cases of COVID-19 is also needed as it is widely claimed that the imposition of the nation-wide lockdown in India on 25 March 2020 has significantly decelerated the progress of COVID-19 pandemic in the country. It has also been claimed that loosening the restrictions under the nation-wide lockdown during its third and the fourth phase has primarily been responsible for the recently witnessed spike in the number of daily reported confirmed cases of COVID-19 in the country. It has even been argued that re-imposing the harsh restrictions as part of the nation-wide lockdown is the only way of stopping or decelerating the progress of COVID-19 pandemic despite the fact that the social and economic cost of nation-wide lockdown has been found to be quite complex and exorbitant. It has repeatedly been stressed that because of serious social and economic implications of the nation-wide lockdown, it cannot be prolonged.

One way of empirically examining these and many other claims is whether the trend in the daily reported confirmed cases of COVID-19 has changed after the imposition of the nation-wide lockdown or after loosening the restrictions under the nation-wide lockdown. If the trend in the daily reported confirmed cases of COVID-19 has changed after the imposition of the nation-wide lockdown, then it can be inferred that the nation-wide lockdown has indeed an impact on the progress of the epidemic. Similarly, if it is found that the trend in the daily reported confirmed cases of COVID-19 has changed after loosening the restrictions under the nation-wide lockdown, then it can be concluded that loosening of the restrictions has been responsible for the spike in reported confirmed COVID-19 cases in the country. However, if there is no change in the trend, then there is little empirical evidence to suggest that either the nation-wide lockdown or loosening of restrictions under the nation-wide lockdown has any telling impact on the progress of the COVID-19 pandemic.

A review of the daily reporting of the confirmed COVID-19 cases in India reveals that during the 28 days from 4 February 2020 through 1 March 2020, no confirmed case of COVID-19 was reported in the country (Table 1). Moreover, during the period 2 March 2020 through 31 March 2020, daily reporting of confirmed COVID-19 cases has been highly inconsistent. For example, no confirmed case of COVID-19 was reported on 3 March; 20 March; and 28 March 2020 whereas on 29 March 2020 alone, 255 confirmed cases of COVID-19 were reported. These inconsistencies in the reporting of daily confirmed cases of COVID-19 may bias any analysis of the trend in the daily reported confirmed cases of COVID-19. It is therefore necessary that the irregular fluctuations in daily reporting of confirmed cases of COVID-19 resulting from the inconsistencies in reporting are ironed out before any analysis of the trend in the reported confirmed cases of COVID-19.

**Table 1:** Reported confirmed cases of COVID-19 in India, 1 March 2020 - 23 May 2020

| Date | Reported confirmed cases | Cumulative confirmed cases | Five days moving average of reported confirmed cases |
| --- | --- | --- | --- |
| January 30, 2020 | 1 | 1 |  |
| January 31, 2020 | 0 | 1 |  |
| February 1, 2020 | 0 | 1 | 1 |
| February 2, 2020 | 1 | 2 | 0 |
| February 3, 2020 | 1 | 3 | 0 |
| February 4, 2020 | 0 | 3 | 0 |
| February 5, 2020 | 0 | 3 | 0 |
| February 6, 2020 | 0 | 3 | 0 |
| February 7, 2020 | 0 | 3 | 0 |
| February 8, 2020 | 0 | 3 | 0 |
| February 9, 2020 | 0 | 3 | 0 |
| February 10, 2020 | 0 | 3 | 0 |
| February 11, 2020 | 0 | 3 | 0 |
| February 12, 2020 | 0 | 3 | 0 |
| February 13, 2020 | 0 | 3 | 0 |
| February 14, 2020 | 0 | 3 | 0 |
| February 15, 2020 | 0 | 3 | 0 |
| February 16, 2020 | 0 | 3 | 0 |
| February 17, 2020 | 0 | 3 | 0 |
| February 18, 2020 | 0 | 3 | 0 |
| February 19, 2020 | 0 | 3 | 0 |
| February 20, 2020 | 0 | 3 | 0 |
| February 21, 2020 | 0 | 3 | 0 |
| February 22, 2020 | 0 | 3 | 0 |
| February 23, 2020 | 0 | 3 | 0 |
| February 24, 2020 | 0 | 3 | 0 |
| February 25, 2020 | 0 | 3 | 0 |
| February 26, 2020 | 0 | 3 | 0 |
| February 27, 2020 | 0 | 3 | 0 |
| February 28, 2020 | 0 | 3 | 0 |
| February 29, 2020 | 0 | 3 | 0 |
| March 1, 2020 | 0 | 3 | 0 |
| March 2, 2020 | 2 | 5 | 5 |
| March 3, 2020 | 0 | 5 | 5 |
| March 4, 2020 | 22 | 27 | 6 |
| March 5, 2020 | 2 | 29 | 6 |
| March 6, 2020 | 2 | 31 | 7 |
| March 7, 2020 | 3 | 34 | 3 |
| March 8, 2020 | 5 | 39 | 4 |
| March 9, 2020 | 5 | 44 | 6 |
| March 10, 2020 | 6 | 50 | 8 |
| March 11, 2020 | 10 | 60 | 8 |
| March 12, 2020 | 14 | 74 | 8 |
| March 13, 2020 | 7 | 81 | 11 |
| March 14, 2020 | 3 | 84 | 11 |
| March 15, 2020 | 23 | 107 | 13 |
| March 16, 2020 | 7 | 114 | 14 |
| March 17, 2020 | 23 | 137 | 22 |
| March 18, 2020 | 14 | 151 | 18 |
| March 19, 2020 | 44 | 195 | 34 |
| March 20, 2020 | 0 | 195 | 41 |
| March 21, 2020 | 88 | 283 | 57 |
| March 22, 2020 | 58 | 341 | 65 |
| March 23, 2020 | 93 | 434 | 82 |
| March 24, 2020 | 85 | 519 | 73 |
| March 25, 2020 | 87 | 606 | 77 |
| March 26, 2020 | 43 | 649 | 58 |
| March 27, 2020 | 75 | 724 | 92 |
| March 28, 2020 | 0 | 724 | 93 |
| March 29, 2020 | 255 | 979 | 120 |
| March 30, 2020 | 92 | 1071 | 182 |
| March 31, 2020 | 180 | 1251 | 248 |
| April 1, 2020 | 385 | 1636 | 264 |
| April 2, 2020 | 329 | 1965 | 366 |
| April 3, 2020 | 336 | 2301 | 425 |
| April 4, 2020 | 601 | 2902 | 486 |
| April 5, 2020 | 472 | 3374 | 491 |
| April 6, 2020 | 693 | 4067 | 595 |
| April 7, 2020 | 354 | 4421 | 593 |
| April 8, 2020 | 853 | 5274 | 608 |
| April 9, 2020 | 591 | 5865 | 676 |
| April 10, 2020 | 547 | 6412 | 787 |
| April 11, 2020 | 1035 | 7447 | 776 |
| April 12, 2020 | 909 | 8356 | 900 |
| April 13, 2020 | 796 | 9152 | 1005 |
| April 14, 2020 | 1211 | 10363 | 987 |
| April 15, 2020 | 1076 | 11439 | 1006 |
| April 16, 2020 | 941 | 12380 | 1045 |
| April 17, 2020 | 1007 | 13387 | 1070 |
| April 18, 2020 | 991 | 14378 | 1165 |
| April 19, 2020 | 1334 | 15712 | 1244 |
| April 20, 2020 | 1553 | 17265 | 1319 |
| April 21, 2020 | 1336 | 18601 | 1403 |
| April 22, 2020 | 1383 | 19984 | 1473 |
| April 23, 2020 | 1409 | 21393 | 1448 |
| April 24, 2020 | 1684 | 23077 | 1579 |
| April 25, 2020 | 1429 | 24506 | 1582 |
| April 26, 2020 | 1990 | 26496 | 1608 |
| April 27, 2020 | 1396 | 27892 | 1651 |
| April 28, 2020 | 1543 | 29435 | 1709 |
| April 29, 2020 | 1897 | 31332 | 1709 |
| April 30, 2020 | 1718 | 33050 | 1889 |
| May 1, 2020 | 1993 | 35043 | 2109 |
| May 2, 2020 | 2293 | 37336 | 2240 |
| May 3, 2020 | 2644 | 39980 | 2677 |
| May 4, 2020 | 2553 | 42533 | 2870 |
| May 5, 2020 | 3900 | 46433 | 3123 |
| May 6, 2020 | 2958 | 49391 | 3272 |
| May 7, 2020 | 3561 | 52952 | 3426 |
| May 8, 2020 | 3390 | 56342 | 3301 |
| May 9, 2020 | 3320 | 59662 | 3552 |
| May 10, 2020 | 3277 | 62939 | 3561 |
| May 11, 2020 | 4213 | 67152 | 3588 |
| May 12, 2020 | 3604 | 70756 | 3668 |
| May 13, 2020 | 3525 | 74281 | 3806 |
| May 14, 2020 | 3722 | 78003 | 3758 |
| May 15, 2020 | 3967 | 81970 | 4034 |
| May 16, 2020 | 3970 | 85940 | 4378 |
| May 17, 2020 | 4987 | 90927 | 4627 |
| May 18, 2020 | 5242 | 96169 | 4956 |
| May 19, 2020 | 4970 | 101139 | 5284 |
| May 20, 2020 | 5611 | 106750 | 5504 |
| May 21, 2020 | 5609 | 112359 | 5768 |
| May 22, 2020 | 6088 | 118447 |  |
| May 23, 2020 | 6564 | 125011 |  |

Source: WHO COVID-19 database

One approach to minimise the impact of reporting inconsistencies in the analysis of the trend in daily reporting of confirmed COVID-19 cases is to use moving average instead of actual daily reported confirmed cases of COVID-19. The same approach has been followed in the present analysis. To minimise the effect of irregular fluctuations in the reporting of COVID-19 cases in the trend analysis, five-day moving average has been used instead of daily reported confirmed cases of COVID-19. In other words, the reported confirmed cased of COVID-19 in a day used in the present analysis are actually the average of the reported confirmed cases of COVID-19 two days prior to the day in question; two days after the day in question; and the reported confirmed cases of COVID-19 on the day in question. For example, the reported confirmed cases of COVID-19 on 3 March 2020 used in the present analysis are actually the simple average of reported confirmed cases of COVID-19 on 1 March through 5 March 2020.

The trend analysis presented in this paper is based on joinpoint regression analysis (Kim et al, 2000). Joinpoint regression analysis is used to study the trend that varies over time. This method first identifies the time point(s) at which the trend in the reported confirmed cases of COVID-19 has changed or the joinpoint(s). Once the joinpoint(s) are identified, then the average per cent change between two joinpoints is calculated to reflect how the trend in the reported confirmed cases of COVID-19 has varied over time. The goal of the joinpoint regression analysis is not to provide the statistical model that best fits the time series data. Rather, the purpose is to provide that model which best summarises the trend in the data (Marrot, 2010). The underlying assumption of joinpoint regression is that trend in the data is not the same throughout the period under reference but is essentially different in different segments of the reference period.

## Joinpoint Regression Model

The joinpoint regression model is essentially different from the conventional piecewise or segmented regression model in the sense that the identification of joinpoint(s) and their location(s) is estimated within the model and are not set arbitrarily as is the case with the piecewise or segmented regression analysis. The minimum and the maximum number of joinpoint(s) are, however, set in advance but the final number of joinpoint(s) or the time point(s) when the trend changes are determined statistically. The model first identifies the time point(s) when there is a significant change in the trend and then calculates the average percentage change *(APC)* which reflects the rate of change between two joinpoint(s). When the number of joinpoint(s) is zero, the model reduces to simple linear regression model.

Let *y_i_* denotes the reported confirmed COVID-19 cases on day *t_i_* such that *t_1_<t_2_<…<t_n_*. Then the joinpoint regression model is defined as

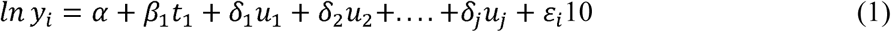

Where

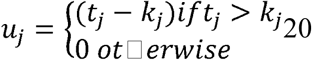

and *k*_1_*<k*_2_…… <*k_j_* are joinpoints. The details of joinpoint regression analysis are given elsewhere (Kim et al, 2000; Kim et al, 2004).

Joinpoint regression analysis has commonly been used when the temporal trend of a given quantity, like incidence, prevalence and mortality is of interest (Tyczynski and Berkel, 2005; Doucet, Rochette and Hamel, 2016; John and Hanke, 2015; Chaurasia, 2020). However, this method has generally been applied with the calendar year as the time scale (Akinyede and Soyemi, 2016; Mogos et al, 2016; Chatenoud et al, 2015; Missikpode at al, 2015). The joinpoint regression analysis can also be applied in epidemiological studies in which the starting date can be easily established such as the day when the disease is detected for the first time as is the case in the present analysis (Rea et al, 2017). Joinpoint regression analysis can, therefore, be applied in the public health framework also and can explore whether the number of reported cases of a specific disease has decreased after the introduction of an intervention to check or control the disease. In the present context, the application of the joinpoint regression analysis can answer the question whether the imposition of the nation-wide lockdown has resulted in the decrease in the reported confirmed cases of COVID-19 in the country or not. If the day of introducing the intervention turns out to be a joinpoint, then, a comparison of daily per cent change in the reported confirmed cases of COVID-19 before and after the joinpoint can tell whether the intervention has been able to bring down the reported confirmed cases of COVID-19 or not.

Actual calculations in the present analysis are carried out using the Joinpoint Regression Program developed by the Statistical Research and Application Branch of the National Cancer Institute of the United States of America (NIC, 2013). The software requires specification of minimum (0) and maximum number of joinpoints (>0) in advance. In the present analysis, the minimum number of joinpoints is specified as 0 while the maximum number of joinpoints have been specified as 5. The programme starts with the minimum number of joinpoints (0, which is actually a straight line and the model is simple linear regression model) and tests whether more joinpoints are statistically significant and must be added to the model (up to the pre-specified maximum number of joinpoints). The tests of significance are based on a Monte Carlo Permutation method (Kim et al, 2000).

This study uses the Bayesian Information Criterion (BIC) to identify the number of joinpoints in the model. The equation for computing the BIC for a *k*-joinpoints model is:

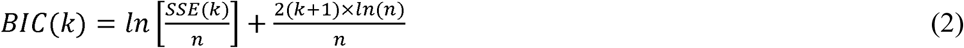

Where SSE is the sum of squared errors of the *k*-joinpoints regression model and *n* is the number of observations. The model which has the minimum value of BIC(*k*) is selected as the final model. There are other methods also for identifying the joinpoints. These include the permutation test method and the data driven BIC methods. Relative merits and demerits of different methods of identifying the joinpoints are discussed elsewhere (NIC, 2013). The permutation test method is regarded as the best method but it is computationally very intensive. It controls the error probability of selecting the wrong model at a certain level (i.e. 0.05). The BIC method, on the other hand, is less computationally intensive.

The Joinpoint Regression Program provides estimates of annual per cent change (*APC*) as it is designed primarily for analysing calendar year data. *APC* is one way to characterise the trend over time. In this approach, a rate or a number is assumed to change at a constant percentage of the rate or the number of the previous year. The rate or the number that changes at a constant percentage every year, changes linearly on a logarithmic scale. In the present case, data on the reported confirmed cases of COVID-19 are available on a daily, not yearly, basis. This means that the *APC* of the Joinpoint Regression Program output is actually the daily per cent change *(DPC)* in the present context. The *DPC* from day *t* to day (*t*+1) is defined as

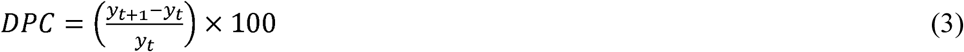

If the trend in the daily reported confirmed cases of COVID-19 is modeled as

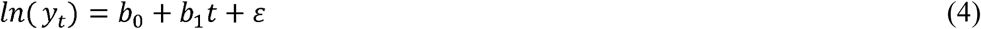

then, it can be shown that the *DPC* is equal to

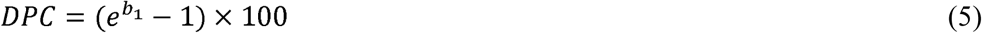

The positive value of *DPC* suggests an increasing trend while the negative value of APC suggests a decreasing trend. The *DPC* reflects the trend in the reported confirmed cases of COVID-19 in different time segments of the reference period identified through joinpoint regression analysis. For the entire reference period, it is possible to estimate average daily percent change *(ADPC)* which is the weighted average of *DPC* in different time segments of the reference period with weights equal to the length of different time segments. However, when the trend changes frequently, *ADPC* has little meaning.

## Trend in Daily Reported Confirmed Cases of COVID-19

Results of the joinpoint regression analysis of the five-days moving average of the daily reported confirmed cases of COVID-19 in India for the period 1 March 2020 through 23 May 2020 are summarised in table 2 and figure 1. The five-days moving average is centred at the mid-point of the five-day interval. For example, the five-day moving average of the period 1 March through 5 March 2020 is centred on 3 March 2020. In other words, the joinpoint regression analysis is carried out for the period 3 March 2020 through 21 May 2020, although, it covers the data on daily reported confirmed cases of COVID-19 from 1 March 2020 through 23 May 2020. The period prior to 1 March 2020 has not been included in the analysis as the daily reported confirmed cases of COVID-19 during the period 30 January 2020 through 1 March 2020 have mostly been found to be zero.

**Figure 1:**
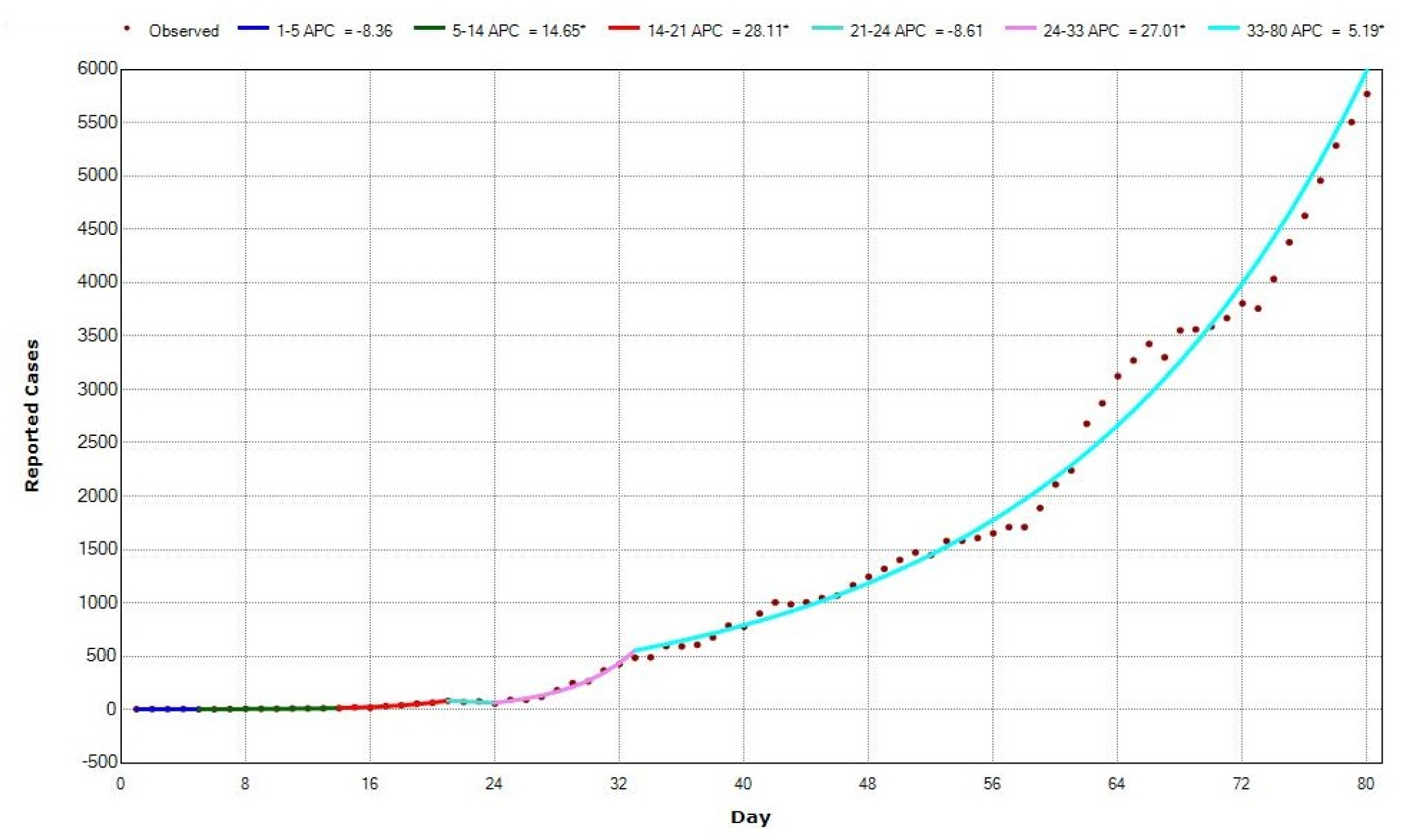
Trend in daily reported confirmed cases of COVID-19 in India during 1 March through 23 May 2020 using joinpoint regression analysis with BIC criterion

**Table 2:** Results of the joinpoint regression analysis

| Segment | Lower<br>Endpoint | Upper<br>Endpoint | Number of<br>days | Daily per<br>cent change<br>(*DPC*) | 95 per cent confidence<br>interval |  | 't' | p>\|t\| |
| --- | --- | --- | --- | --- | --- | --- | --- | --- |
|  |  |  |  |  | Lower | Upper |  |  |
| 1 | 3 March 2020 | 7 March 2020 | 5 | -8.363 | -16.063 | 0.044 | -1.988 | 0.051 |
| 2 | 7 March 2020 | 16 March 2020 | 10 | 14.648* | 11.228 | 18.173 | 9.021 | 0.000 |
| 3 | 16 March 2020 | 23 March 2020 | 8 | 28.113* | 22.242 | 34.267 | 10.553 | 0.000 |
| 4 | 23 March 2020 | 26 March 2020 | 4 | -8.612 | -30.761 | 20.622 | -0.648 | 0.519 |
| 5 | 26 March 2020 | 4 April 2020 | 10 | 27.008* | 23.219 | 30.913 | 15.777 | 0.000 |
| 6 | 4 April 2020 | 21 May 2020 | 48 | 5.194* | 4.972 | 5.416 | 47.944 | 0.000 |
| All | 3 March 2020 | 21 May 2020 | 80 | 9.094<sup>#</sup> | 7.694 | 10.513 | 13.206 | 0.000 |

Source: Author's calculations
Remarks: \* Statistically significant.
<sup>#</sup> Average daily per cent change (*ADPC*)
Number of days in a segment include both lower endpoint and upper endpoint.

The application of the joinpoint regression analysis divides the duration 1 March 2020 through 23 May 2020 or a period of 84 days into five-time segments and the trend in the daily reported confirmed cases of COVID-19 is found to be different in different time segments. During the first five days of the period under reference - 3 March 2020 (day 1) through 7 March 2020 (day 5), the trend in the daily reported confirmed cases of COVID-19 in the country has been found to be negative which means that daily reported confirmed cases of COVID-19 in the country actually decreased, instead increased, during this period at a daily per cent decrease *(DPC)* of around 8 per cent per day. This decrease in the reported confirmed cases of COVID-19 may be attributed to reporting inconsistencies, although the *DPC* during this period has been found to be statistically significant. On the other hand, during the next 10 days - from 7 March 2020 (day 5) through 16 March 2020 (day 14) - the daily reported confirmed cases of COVID-19 increased daily per cent increase *(DPC)* of almost 15 per cent. The increase in the daily reported confirmed cases of COVID-19 accelerated further during the next eight days - from 16 March 2020 (day 14) through 23 March 2020 (day 21) - when the daily reported confirmed cases of COVID-19 in the country increased at a *DPC* of more than 28 per cent. However, during the next five days - between 23 March 2020 (day 21) through 26 March 2020 (day 24) - the reported confirmed cases of COVID-19 decreased again with a *DPC* of almost −9 per cent, although the daily per cent decrease in during this period has not been found to be statistically significant. The daily reported confirmed cases of COVID-19 increased again with a *DPC* of around 27 per cent during the next 10 days - from 26 March 2020 (day 24) through 4 April 2020 (day 33). After 4 April 2020, however, there has been no change in the trend in the daily reported number of confirmed cases of COVID-19 till 21 May 2020. During the 43 days from 4 April 2020 through 21 May 2020, the daily reported confirmed cases of COVID-19 in the country increased at a *DPC* of almost 5.2 per cent.

It is also possible to estimate the average daily per cent change (*ADPC*) in the reported confirmed cases of COVID-19 in the country during the period 3 March 2020 through 21 2020. The joinpoint regression analysis suggests that the reported confirmed cases of COVID-19 in the country increased at an *ADPC* of slightly more than 9 per cent during this period (Table 3). The *APDC*, however, has little relevance in the present case as the trend in the reported confirmed cases of COVID-19 during this period has been quite erratic, particularly during the period 3 March 2020 through 4 April 2020. After 4 April 2020, however, there is no change in the trend and, therefore, the *ADPC* is the same as *APC* during this period.

**Table 3:** Forecast of daily reported confirmed cases of COVID-19 till 15 June 2020

| Date | Estimate | 95% Confidence interval |  |
| --- | --- | --- | --- |
|  |  | Lower | Upper |
| May 22, 2020 | 6301 | 5694 | 6972 |
| May 23, 2020 | 6629 | 5977 | 7350 |
| May 24, 2020 | 6973 | 6274 | 7748 |
| May 25, 2020 | 7335 | 6586 | 8168 |
| May 26, 2020 | 7716 | 6913 | 8610 |
| May 27, 2020 | 8117 | 7257 | 9076 |
| May 28, 2020 | 8539 | 7618 | 9568 |
| May 29, 2020 | 8982 | 7997 | 10086 |
| May 30, 2020 | 9449 | 8394 | 10633 |
| May 31, 2020 | 9939 | 8812 | 11208 |
| June 1, 2020 | 10456 | 9250 | 11815 |
| June 2, 2020 | 10999 | 9710 | 12455 |
| June 3, 2020 | 11570 | 10192 | 13130 |
| June 4, 2020 | 12171 | 10699 | 13841 |
| June 5, 2020 | 12803 | 11231 | 14591 |
| June 6, 2020 | 13468 | 11790 | 15381 |
| June 7, 2020 | 14167 | 12376 | 16214 |
| June 8, 2020 | 14903 | 12991 | 17092 |
| June 9, 2020 | 15677 | 13637 | 18018 |
| June 10, 2020 | 16492 | 14315 | 18994 |
| June 11, 2020 | 17348 | 15027 | 20022 |
| June 12, 2020 | 18249 | 15774 | 21107 |
| June 13, 2020 | 19197 | 16558 | 22250 |
| June 14, 2020 | 20194 | 17381 | 23455 |
| June 15, 2020 | 21243 | 18246 | 24725 |

Source: Author's calculations

The joinpoint regression analysis suggests that the trend in the daily reported confirmed cases of COVID-19 changed statistically significantly at the 5^th^ day (7 March 2020); 14^th^ day (16 March 2020); 24^th^ day (26 March 2020); and 33^rd^ day (4 May 2020) of the period beginning 3 March 2020. The nation-wide lockdown in the country was imposed on 25 March 2020 initially for a period of 21 days which was then extended to 3 May 2020 on 15 April 2020. On 4 May 2020, the lockdown was again extended up to 17 May 2020 but with a relaxed set of restrictions which, on 18 May 2020, was further extended to 31 May 2020 with even more relaxed set of restrictions. The present analysis suggests that the day of the change in the trend in daily reported confirmed cases of COVID-19 in the country has never matched with the date of imposing the lockdown or dates of first, second and third extensions of the lockdown. There has also been no change in the trend in the daily reported confirmed cases of COVID-19 after 4 April 2020. If the change in the trend is any indication of the effectiveness of the interventions directed towards changing the course of the pandemic, then, it is obvious from the present analysis that, neither the lockdown imposed on 25 March 2020 nor the three extensions of the lockdown appears to have resulted in any significant change in the trend in the reported confirmed cases of COVID-19 in the country. In other words, it may be concluded that the strategy of the nation-wide lockdown has not been effective in changing the course of the COVID-19 pandemic in the country.

The joinpoint regression analysis also suggests that the trend in the reported confirmed cases of COVID-19 has been different during the period 4 April 2020 onwards as compared to the trend prior to 4 April 2020. Before 4 April 2020, the trend in the reported confirmed cases of COVID-19 in the country has been very erratic and is characterised by change in the direction of the trend in a very short duration. There is no apparent reason for the observed frequent changes in the direction of the trend in the reported confirmed cases of COVID-19 and that too in a short to very short duration. It therefore appears that the observed frequent changes in the trend in a very short duration in the reported confirmed cases of COVID-19 in the country prior to 4 April 2020 are essentially the result of the inconsistencies in the reporting of COVID-19 cases. It is only after 4 April 2020 that the consistency in the daily reporting of the confirmed cases of COVID-19 appears to have improved. Because of the highly erratic nature of the trend in the reported confirmed cases of COVID-19 prior to 4 April 2020, any inference about the progress of the COVID-19 pandemic on the basis of the trend during any period before 4 April 2020 should be desisted and the interpretations about the progress of the pandemic on the basis of the trend in the reported confirmed cases of COVID-19 prior to 4 April 2020 should be made with caution. The consistency of the trend in the reported confirmed cases of COVID-19 after 4 April 2020 suggests that any inference about the progress of the COVID-19 pandemic in the country should be made on the basis of the trend in the reported confirmed cases of COVID-19 during the period 4 April 2020 through 21 May 2020 only which shows that there has been no smooth change in the progress of the COVID-19 pandemic in the country since 4 April 2020.

## Forecasting Number of COVID-19 Cases

The *DPC* in the daily reported confirmed cases of COVID-19 during the period 4 April 2020 through 21 May 2020 may be used for forecasting the daily reported confirmed cases of COVID-19 in the immediate future under the assumption that the trend in the daily reported confirmed cases of COVID-19 remains unchanged. This exercise suggests that by 15 June 2020, the daily reported confirmed cases of COVID-19 in the country is likely to increase to 21243 with a 95 per cent confidence interval of 18246 - 24725 (Table 3 and Figure 2). This increase in the daily reported confirmed cases of COVID-19 may change only when there is a significant change in the trend which is possible only when an appropriate set of new interventions are introduced to combat COVID-19 pandemic in the country. The trend during 4 April 2020 through 21 May 20202 in the reported confirmed cases of COVID-19 in the country suggests that the nation-wide lockdown imposed on 25 March 2020 and extended three times subsequently has largely been irrelevant in checking the progress of the COVID- 19 pandemic because of a host of factors. It was expected that the nation-wide lockdown will be able to break the transmission of the novel coronavirus which causes COVID-19. However, the national-wide lockdown could not check large scale movement, especially of migrant workers from urban areas to rural hinterland which appears to be largely responsible for the increase in the reported confirmed cases of COVID-19 in the country. In order to change the course of the pandemic, it is, therefore, imperative that efforts beyond the lockdown should be undertaken to modify the course of the pandemic. One such intervention is the population-wide testing for COVID-19 followed by isolation of the positive cases and their contacts. The need for such a strategy also stems from the fact that a very large proportion of the individuals tested positive for COVID-19 have been found to be asymptomatic. Chaurasia (2020a) has suggested a cluster-based approach of population-wide testing for COVID-19 in the country which significantly reduces the number of tests to be done.

**Figure 2:**
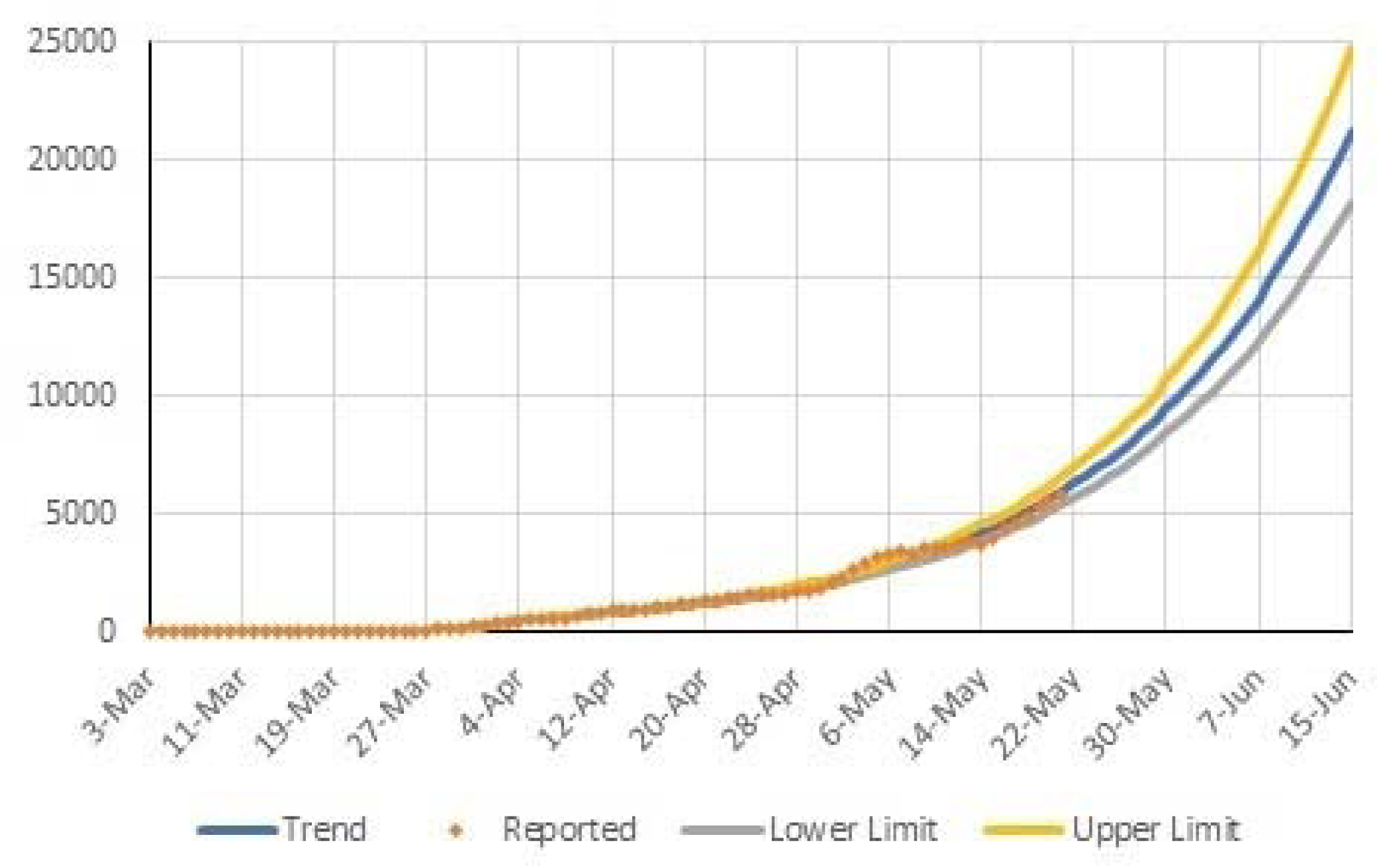
Forecast of daily reported confirmed COVID-19 cases in India up to 15 June 2020

The forecast of the daily reported confirmed cases of COVID-19, on the basis of the joinpoint regression analysis also suggests that the total number of confirmed COVID-19 cases in the country are likely to increase to almost 422 thousand by 15 June 2020 with a 95 per cent confidence interval ranging from around 376 thousand to around 473 thousand if the present trend in the daily reported confirmed cases of COVID-19 continues in the days to come. This implies that the daily reported confirmed cases of COVID-19 in the country are bound to increase quite rapidly in the days to come. This forecast about the increase in the reported confirmed cases of COVID-19 in the country in the days to come can be changed only through the introduction of appropriate interventions.

## Conclusions

The present analysis, based on the daily reported confirmed cases of COVID-19, suggests that there has virtually been little impact of the nation-wide lockdown and its subsequent extensions and relaxations in restrictions on the progress of the COVID-19 pandemic in India. There has also been little empirical evidence to suggest that relaxations in the restrictions under the third and the fourth extension of the nation-wide lockdown have resulted in spiking of the reported confirmed cases of COVID-19 in the country as is generally claimed. This conclusion is based on the hypothesis that if the nation-wide lockdown and extensions that followed have been effective, the trend in the reported confirmed cases of COVID-19 in the country would have changed statistically significantly. The present analysis suggests that any change in the trend in the reported confirmed cases of COVID-19 appears to be largely the result of the inconsistencies in the reporting of COVID- 19 cases.

Based on the trend in the reported confirmed cases of COVID-19 during the period 4 April 2020 through 21 May 2020, the analysis also suggests that the daily reported confirmed cases of COVID-19 in the country are likely to increase to somewhere between 18246 to 24726 with an average of around 21 thousand by 15 June 2020 whereas the total number of confirmed cases of COVID-19 are the most likely to increase to somewhere between around 422 thousand with a 95 per cent confidence interval of 376 to 473 thousand by 15 June 2020. Appropriate interventions, in addition to the nation-wide lockdown, are needed to change this trend in the progress of the COVID-19 pandemic in the country. One such intervention that may contribute to modifying the progress of the pandemic in the country may be population-wide testing for COVID-19 along with isolation of positive cases and quarantine of contacts to the identified positive cases.

## Data Availability

Data table is included in the paper itself.

